# Effects of ultrasound-guided high ankle block combined with general anesthesia on postoperative cognitive function in fragile elderly patients based on the analysis of scale and EEG monitor

**DOI:** 10.1101/2024.11.02.24316632

**Authors:** Ziwei Xia, Guangkuo Ma, Huanjia Xue, Fangjun Wang, Liwei Wang, Kai Wang

**Author notes:** **Corresponding author:** Kai Wang.

## Abstract

**Introduction:** Ultrasound-guided high ankle block can provide prolonged analgesia for foot and ankle surgeries while preserving ankle motor function. Given that fragile elderly patients are prone to cognitive impairment after general anesthesia, this study intends to implement high ankle block for fragile elderly patients undergoing foot and ankle fracture surgery, and use intraoperative electroencephalogram (EEG) analysis combined with postoperative scales to investigate its effect on patients’ postoperative cognitive function.

**Methods and analysis:** This randomized controlled trial will be conducted in Xuzhou Central Hospital. A total of 126 elderly patients scheduled to undergo unilateral ankle fracture (uni/bi-/triple ankle) incision and reduction internal fixation (ORIF) surgery will be randomly assigned to either the HAB group or the GB group in a 1:1 ratio. Patients will receive ultrasound-guided high ankle block and general anesthesia (HAB group) or general anesthesia alone (GB group). Multimodal analgesia will include intraoperative flurbiprofen ester and postoperative patients will be given appropriate medications according to the postoperative pain management “three-step ladder”. The primary outcome indicator was the incidence of dNCR at 7th day postoperation. The secondary outcome indicators were the incidence of dNCR at 1st and 30th days postoperation; alpha relative power, alpha power, and burst inhibition ratio in each brain region at 30 min after induction of anesthesia; MoCA scores, patient stress response indexes: ACTH, NE, Ang-II, β-EP, Glu, and Cor levels; patients’ VAS pain scores; patients’ satisfaction scores; first time getting out of bed postoperatively; and time of discharge from the hospital. Safety outcomes were dizziness, headache, vomiting, urinary retention, bradycardia, tachycardia, hypotension, and hypertension.

**Ethics and dissemination:** Ethics approval was obtained from the Ethics Committee of the Xuzhou Central Hospital. All patients will provide written informed consent. The results of this study will be published in a peer-reviewed journal.

**Trial registration number:** Chinese Clinical Trial Registry (ChiCTR2400050927)

## Introduction

The ankle is the largest weight-bearing joint in the human body, and ankle fractures top the list of intra-articular fractures. Accounting for 55.7% of all foot and ankle fractures, ankle fractures are the most common type of lower extremity fracture and one of the most common types of fractures worldwide [1]. And with the progress of aging population, more and more elderly people need to undergo unilateral ankle fracture (uni/bi-/triple ankle) ORIF surgery. And Perioperative Neurocognitive Disorders (PND), which encompasses delayed neurocognitive recovery (dNCR), is a common central nervous system complication after surgery, which is mainly manifested by the emergence of postoperative decline in cognitive functions such as learning, memory, mood, emotion and judgment. It has been found that cognitive dysfunction mainly occurs in general surgery, orthopedics, and some upper abdominal surgeries [2, 3], of which the prevalence rate is generally 6.7% in people aged 60-64 years, while the prevalence rate in people aged 80 years or older is 25.2% [4]. And previous studies have shown that fragile patients have a high incidence of perioperative cognitive dysfunction [5]. Therefore, how to reduce the occurrence of perioperative cognitive dysfunction in fragile elderly patients with unilateral ankle fracture (single/biple/triple ankle) ORIF is one of the hotspots of clinical research at present.

Intravertebral anesthesia is often used for lower limb fracture surgery, but elderly patients are often associated with lumbar disc herniation, ligament calcification and other problems, and are prone to urinary retention, postoperative headache and other adverse reactions, which makes it impossible to carry out intravertebral anesthesia. Simple general anesthesia due to the gradual deterioration of the function of various organs in elderly patients, reduced immunity, vascular elasticity and poor tolerance to anesthesia, coupled with the traumatic stimulation of the operation, which often causes significant stress reactions, and may even lead to perioperative cognitive dysfunction, which is not conducive to the prognosis of the patient [6]. Some studies have shown that general anesthesia combined with femoral and sciatic nerve block is more capable of promoting hemodynamic stability, providing a favorable environment for the smooth implementation of surgery. The reason may be that ropivacaine can effectively inhibit the transmission of surgical injury stimuli to the central nervous system, while femoral and sciatic nerve block can precisely inject local anesthetic drugs into the target plexus; coupled with general anesthesia to inhibit the hypothalamus and the limbic system of the cerebral cortex of the projection system, the two anesthesia programs from different mechanisms of action to reduce the surgical stimuli to stabilize the hemodynamics of the body[7, 8]. The nerves innervating sensation and movement in the foot and ankle are branches of the femoral and sciatic nerves, mainly including the tibial, deep peroneal, superficial peroneal, peroneal and saphenous nerves [9]. Blocking five nerves in the plane of the ankle joint and below (classic ankle block) provides perfect analgesia for surgery of the forefoot [10]. Based on this, Klaus et al. [11] proposed the concept of “high ankle block” in 2022, which provided perfect analgesia for three patients undergoing ankle surgery by moving the block to about 15 cm proximal to the ankle joint and successfully preserved the motor function of the lower leg. Therefore, for orthopedic foot and ankle surgeries, blocking the tibial, superficial peroneal and deep peroneal nerves, peroneal nerve and saphenous nerve in the middle of the calf between the popliteal fossa and the ankle may be able to provide prolonged analgesia while preserving the motor function of the ankle joint.

Scale assessments have been commonly used in the past to assess postoperative cognitive dysfunction, and they provide a standardized method for assessing a patient’s cognitive function, making the results comparable and objective. Most cognitive function scales are simple in design and easy to implement quickly in a clinical setting. However, the use of scales to assess postoperative cognitive function lacks a gold standard, and different scales may focus on different cognitive domains, leading to differences in assessment results. Therefore, in this study, the Z-value composite score was used, and a total of 7 scales were included to comprehensively assess postoperative cognitive function. Intraoperative electroencephalogram was further included to assess postoperative cognitive function more objectively.

EEG provides physiologic information about a patient’s brain electrical activity and is a noninvasive test with high temporal resolution. EEG was first used to monitor the brain state of patients with psychiatric disorders with a view to deriving the underlying neuropathological mechanisms of psychiatric disorders, Lucey et al. found that reduced NREM SWA was associated with poor cognitive performance and Alzheimer’s disease pathology [12]. Zheng Liyun et al. found that event-related potential studies confirmed the presence of attentional bias as well as abnormalities in inhibitory control in patients with eating disorders (ED) at the brain electrophysiological level [13]. With the development of the brain-computer interface field, the availability and portability of EEG devices have further facilitated the use of EEG in perioperative neurocognitive dysfunction [14, 15].Melody Reese et al. found that burst inhibition and preoperative cognitive deficits were associated with perioperative neurocognitive dysfunction [16]; the correlation between PND and the power of the EEG alpha frequency band. The EEG alpha frequency band originates in the thalamus and is involved in the regulation of arousal, attention and other important cognitive functions. During the anesthesia maintenance phase, the EEG power of all frequency bands decreases with age, with the most obvious change in the α-band power. Therefore, the study of α-band power has become a hot spot for PND analysis. Meanwhile, several previous studies have shown that frontal α power and burst inhibition ratio under general anesthesia were found to be related to aging and cognitive function reduction [17, 18]. For example, in patients anesthetized with propofol and sevoflurane, the incidence of burst suppression increased from 0 to 1 as the α-band power decreased from >15 dB to <-5 dB during induction, and the incidence of PND increased accordingly, which may be an objective indication of a “fragile brain” [19]. In addition, intraoperative α-band power was reduced in patients with PND and was independent of the anesthetic dose[20], which may be related to the disruption of thalamocortical feedback mechanisms related to intraoperative α-band power in patients with PND. Occipital alpha relative power under general anesthesia was later found to be associated with the onset of delirium both with eyes open or closed, and occipital alpha relative power was negatively correlated with the severity of delirium in the eyes closed state [21].

In summary, it is proposed to implement high ankle block and general anesthesia for elderly unilateral ankle fracture (single/biple/triple ankle) ORIF surgery patients to determine the incidence of delayed neurocognitive recovery obtained from the neuropsychological test scale question group and record the whole-brain α-relative power, α-power, and burst-inhibition ratio to explore its effect on postoperative cognitive function in vulnerable elderly patients, so as to reduce the incidence of such patients’ postoperative cognitive This will reduce the occurrence of postoperative cognitive dysfunction in these patients, thereby improving the quality of postoperative recovery and enabling patients to recover early.

### Objectives

The aim of this study was to compare the effects of ultrasound-guided high ankle block + general anesthesia versus general anesthesia alone in fragile elderly patients undergoing unilateral ankle fracture (uni/bi-/triple ankle) incision and reduction internal fixation (ORIF) surgery. We hypothesized that an ultrasound-guided high ankle block and general anesthesia regimen would reduce the incidence of dNCR and improve postoperative recovery.

## Methods and analysis

Reporting for this program follows the guidelines of the Standard Program Item: Recommendations for Interventional Trials (SPIRIT)

### Study design and patients

This is a single-center, prospective, randomized, patient-blinded and assessment-blinded, parallel-group controlled, superiority clinical trial. A total of 126 patients will be enrolled in Xuzhou City Central Hospital. Xuzhou City Central Hospital is a tertiary teaching hospital. We plan to enroll patients between September 1, 2024 and December 30, 2025. The study flow chart is shown in **Figure 1**.

**Figure 1.**
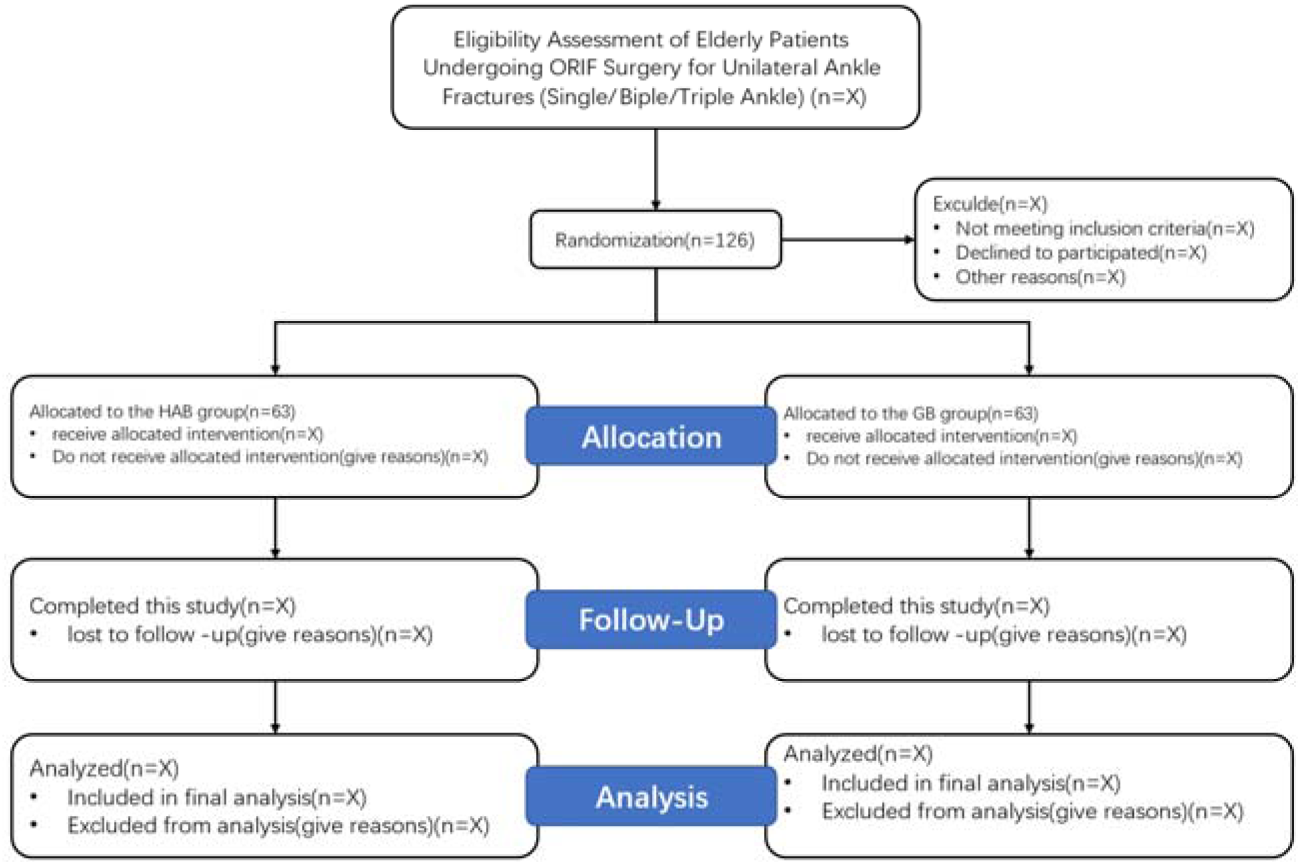
Study flow diagram.

### Inclusion criteria

1. Age ≥ 60 years old, good communication skills and can cooperate with the completion of various testing operations;
2. American Society of Anesthesiologists (ASA) grade I-III;
3. Preoperative Mental State Examination Scale (MMSE) score, the normal cut-off value of the criteria: illiterate (uneducated) group > 17 points, elementary school (≤ 6 years of education) group > 20 points, secondary school or above (> 6 years of education) group > 24 points;
4. Patients eligible for fragile brain function;
5. Patients who underwent unilateral ankle fracture (uni/bi-/triple ankle) incision and reduction internal fixation (ORIF) surgery.

### Exclusion criteria

1. BMI ≥40kg/m^2^;
2. History of allergy to local anesthetic and contraindication to nerve block anesthesia;
3. Those who suffer from serious heart, liver, lung, kidney and other vital organ diseases;
4. People with serious infection at the puncture site and obvious abnormality of coagulation function;
5. Cognitive or mental dysfunction resulting in the inability to communicate verbally for pain scoring;
6. Do not agree to participate in this study or sign the informed consent.

### Rejection criteria

1. Voluntary withdrawal;
2. Failure of follow-up;
3. Subject’s disobedience to the arrangement or agreement of the trial;
4. Acceptance of other analgesic treatment regimens during follow-up.

### Randomisation and blinding

An independent investigator will use an online tool to generate random numbers (https://www.sealedenvelope.com/simple-randomiser/v1/lists) in a 1:1 allocation ratio, and the randomized results will be stored in sealed opaque envelopes. Patients will be randomly assigned to either the HAB group or the GB control group with general anesthesia alone (63 in each group). Due to differences in anesthesia methods, it will not be possible to blind the patients, or the anesthesiologists to the groups; however, they will not be involved in patient recruitment, data collection, or statistical analysis. Subgroup blinding was performed for surgeons, postoperative care providers, outcome assessors, and the person responsible for statistical analysis.

### Study interventions

15 minutes before the induction of anesthesia in the HAB group, a high ankle block on the affected side was performed for the HAB group: a. The long axis of the ultrasound probe was perpendicular to the longitudinal axis of the body, and was placed in the anterior lateral aspect of the tibiofibular bone of the calf, about 15 cm from the external ankle. The deep peroneal and superficial peroneal nerves were blocked with 10 ml each of 0.375% ropivacaine. b. The long axis of the ultrasound probe was perpendicular to the longitudinal axis of the body, and placed at the posterior lateral side of the calf at a distance of about 15 cm from the external ankle. The tibial nerve was blocked using 10 ml of 0.375% ropivacaine. c. The long axis of the ultrasound probe was perpendicular to the longitudinal axis of the body, placed posterior to the lateral ankle, and shifted upward to 15 cm from the lateral ankle. The peroneal nerve was blocked using 5 ml of 0.375% ropivacaine; d. The ultrasound probe was placed perpendicular to the longitudinal axis of the body, anterior to the medial ankle, and moved up to 15 cm from the medial ankle. The saphenous nerve was blocked using 0.375% ropivacaine 5 ml. General anesthesia induction: the same induction protocol was used in both groups, and the same induction protocol was used (midazolam 0.05 mg/kg, etomidate 0.3 mg/kg, sufentanil 0.5 μg/kg, rocuronium bromide 0.6 mg/kg). The patient’s consciousness disappeared, the muscle relaxation was placed into the endotracheal tube, and the respiratory parameters were set as follows: inhalation oxygen flow rate of 2 L/min, tidal volume of 6-8 ml/kg, respiratory rate of 10-14 breaths/min, inhalation/exhalation ratio of 1:1.5, and maintenance of PETCO of 235-45 mmHg. Anesthesia maintenance: the HAB group was maintained with static-aspiration combined anesthesia, remifentanil (0.1ug/kg/min), Propofol (4-6mg/kg/h) and 0.8-1.2% sevoflurane by inhalation. And the GB group was maintained with sedation combined anesthesia with remifentanil (0.3ug/kg/min), propofol (4-6mg/kg/h), and inhalation of 0.8-1.2% sevoflurane. The timeline for patient enrollment, study interventions, and outcome assessment was in accordance with the SPIRIT statement (**Table 1**).

**Table 1.**
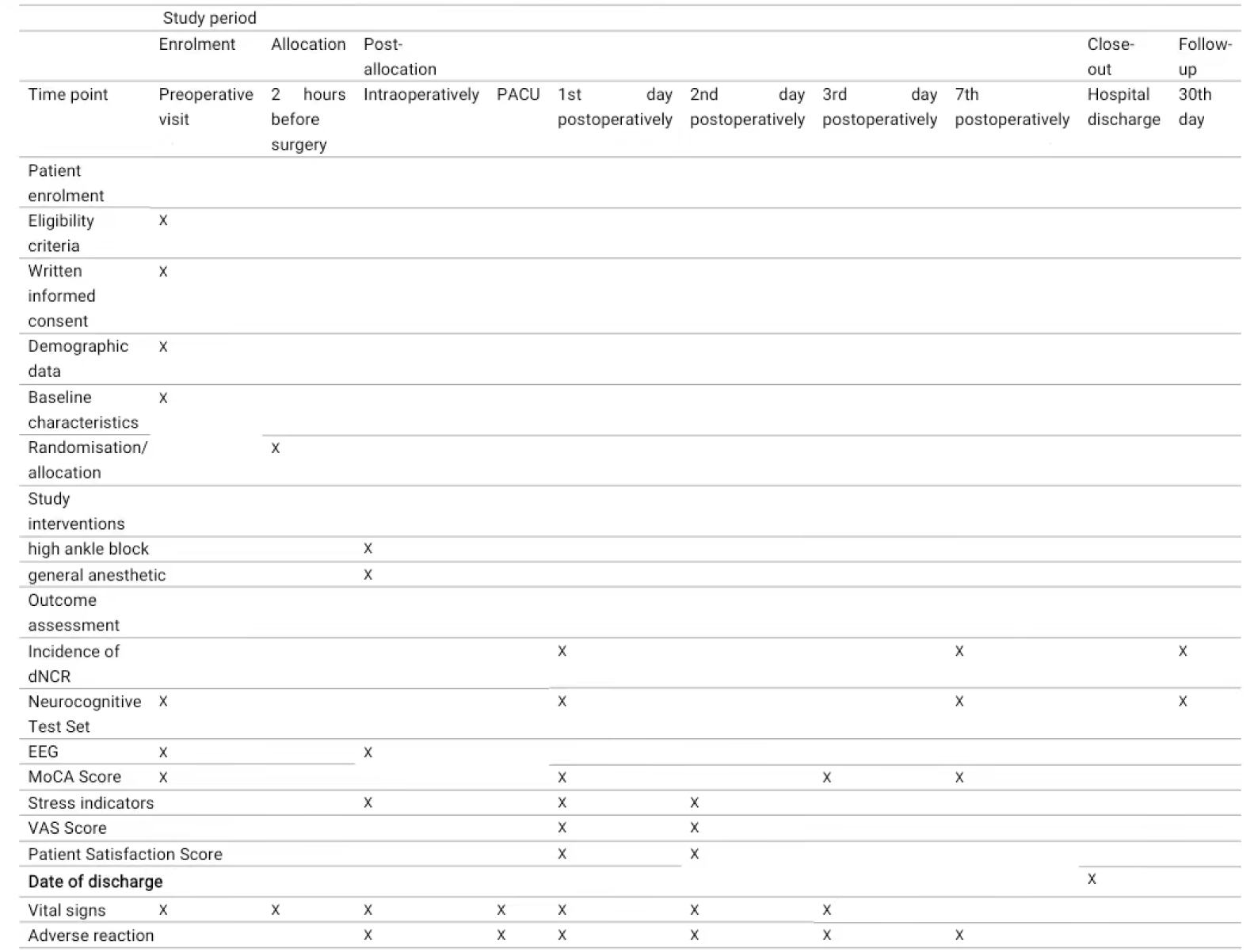
Schedule of patient enrollment, study interventions and outcome assessment.

### Anaesthetic care

The patient will be preoperatively fasted for 6-8 hours and have no premedication. A preoperative EEG will be obtained from the patient in the ward. In the operating room, patients will have their baseline blood pressure and heart rate measured prior to anesthesia and will receive ECG, noninvasive blood pressure, pulse oximetry (SpO_2_) and EEG.

For the HAB group a high ankle block on the affected side was performed 15 min before induction of anesthesia. The same induction regimen (midazolam 0.05 mg/kg, etomidate 0.3 mg/kg, sufentanil 0.5 μg/kg, rocuronium bromide 0.6 mg/kg) was given. The patient’s consciousness disappeared, the muscle relaxation was placed into the tracheal tube, and the respiratory parameters were set as follows: inhalation oxygen flow rate of 2 L/min, tidal volume of 6-8 ml/kg, respiratory rate of 10-14 breaths/min, and inhalation/exhalation ratio of 1:1.5. After anesthesia was induced, radial artery puncture was performed, and the arterial blood pressure was monitored.

During anesthesia maintenance: remifentanil (0.1ug/kg/min) was used in the HAB group, while remifentanil (0.3ug/kg/min) was used in the GB group. A suitable depth of anesthesia was maintained by adjusting the concentration of inhaled sevoflurane. The surgical volume tracing index (SPI) was also maintained between 20-50 indicating a suitable level of analgesia. To ensure intraoperative muscle relaxation, patients will receive additional administration of rocuronium bromide. Patients will also be given an intravenous infusion of lactated Ringer’s solution intraoperatively. After surgery, all patients will be transferred to the anesthesia recovery unit (PACU).

The Modified Observer Assessment of Alertness and Sedation Scale (MOAA/S) and Modified Aldrete Score will be assessed every 5 minutes. A modified Aldrete score ≥9 indicated readiness for PACU discharge to the surgical ward

For analgesia, patients in both groups were given intravenous flurbiprofenate 50 mg 30 min before the end of surgery to assist with analgesia, and all anesthetic drugs were discontinued 5 min before the end of surgery. Postoperative pain will be assessed using a numerical rating scale ranging from 0 to 10 (0=no pain, 10=worst pain imaginable). If the patient complains of a score >4, medication will be given by the ward physician in accordance with the “three-step ladder” of postoperative pain management. The time of the first remedial analgesia, the number of remedial analgesia and the amount of analgesic drugs used within 72 h, and the remedial analgesia rate were recorded.

### Primary and secondary outcomes

The primary outcome of this trial is the incidence of dNCR at 7 days postoperatively. We will assess postoperative cognitive dysfunction with postoperative neuropsychological test question sets and intraoperative EEG changes. The secondary endpoints are the incidence of dNCR at 1 and 30 days postoperatively; alpha relative power, alpha power, and burst inhibition ratio in each brain region at 30 min after induction of anesthesia; MoCA scores, patient stress indices: levels of ACTH, NE, Ang-II, β -EP, Glu, and Cor; patients’ VAS pain scores; patients’ satisfaction scores; time to first time out of bed postoperatively; and time to discharge from the hospital.

Neuropsychological test combination tests were performed at 1, 7, and 30 days postoperatively, and the incidence of dNCR was recorded; alpha relative power, alpha power, and burst inhibition ratio of each brain region were recorded at 30 min after induction of anesthesia; MoCA scores were recorded at 3 and 7 days postoperatively; intraoperative, postoperative day 1, and postoperative day 2 patients’ indicators of stress response: ACTH, NE, Ang-II, β-EP, Glu, and Cor levels ; patients’ VAS pain scores on postoperative day 1 and postoperative day 2; patients’ satisfaction scores on postoperative day 1 and postoperative day 2; first time out of bed and time of discharge can be recorded during postoperative telephone follow-up.

### Safety outcomes

Including perioperative dizziness, headache, vomiting, urinary retention, bradycardia, tachycardia, hypotension, and hypertension, the patients’ mean arterial pressure (MAP) and heart rate (HR) were recorded as the basal blood pressure and heart rate after 5 min of admission to the room, and the occurrence of a decrease in MAP of more than 30% of the basal value within 30 min after anesthesia was considered to be hypotension, and ephedrine 5 mg was given intravenously, and the occurrence of hypotension within 30 min after anesthesia was recorded. Ephedrine 5 mg was administered intravenously, and the occurrence of hypotension within 30 min after anesthesia was recorded. Bradycardia was defined as HR <45 beats/min for at least 1 minute; hypertension was defined as an increase in mean blood pressure >30% of baseline for at least 1 minute; and tachycardia was defined as HR >100 beats/min for at least 1 minute. These hemodynamic events will be assessed during anesthesia and in the PACU, and interventions will be at the discretion of the attending anesthesiologist using atropine 0.3-0.5 mg or uradil 5 m. Episodes of headache, dizziness, nausea and vomiting will be documented for the first 48 hours postoperatively.

### Data collection and monitoring

Independent researchers will collect demographic data (age, gender, height, weight, and BMI), baseline characteristics (preoperative medications, comorbidities, ASA classification, smoking status, level of education, 10min EEG signals at rest with eyes closed for 1 day prior to surgery with MMSE scores, MoCA scores, and perioperative data (alpha relative power in each brain region, alpha power, burst suppression ratio, MMSE score, dNCR incidence, patient stress indicators: ACTH, NE, Ang-II, β-EP, Glu, Cor levels, VAS pain scores, patient satisfaction scores, first time out of bed, and discharge time);

All data will be collected on a case report form and then entered into an electronic database under the supervision of another researcher. An independent Data Monitoring Committee (DMC) will provide ongoing review of data collection. The electronic database will be locked once data registration is complete. Data sets without personally identifiable information will be sent to independent statisticians for final analysis according to a pre-specified statistical plan.

### Sample size calculation

PASS 2015 software was used for sample size calculation. The primary observation in this study was the incidence of postoperative 7-day dNCR. In previous studies, the incidence of postoperative 7-day dNCR in patients undergoing foot and ankle surgery was approximately 35%. Assuming that the improvement rate of ultrasound-guided high ankle block is 65%, with the same number of people in both groups K=1, the same standard deviation σ, a test level α of 0.05, and a test efficacy β of 80%, a two-sided test was performed, and the minimum sample size was calculated to be 50 cases, and the final sample size for each group was determined to be 63 cases, taking into account the 20% shedding rate.

### Statistical analysis

SPSS 24.0 and SPSS.0 Prism 8.0 were used for statistics and plotting. Measurement information was determined to follow normal distribution using the Shapiro-Wilk test; measures conforming to normal distribution were expressed as mean ± standard deviation (x ± s), two independent samples t-test was used for comparison between groups, and repeated measures information was analyzed using the repeated measures ANOVA; non-normally distributed measures were expressed as median and interquartile spacing using the Mann-Whitney U test. Count data were expressed as rates; the chi-square test or Fisher’s exact probability method was used. Comparison of rank information was performed using the multiple-sample rank sum test. The test level α was taken as 0.05, and two-sided tests were used for all analyses, with differences considered statistically significant at P < 0.05. Interim analyses were not planned and missing data will not be entered.

### Patient and public involvement

Patients and the public are not involved in the design, recruitment, conduct or reporting of the study, design, recruitment, conduct, or reporting of the study. The final study results will be e-mailed to participants.

### Ethics and dissemination

This study was approved by Xuzhou Central Hospital Biomedical Research Review Committee on 12 September 2024. The protocol was registered in the China Clinical Trial Registry on 27 September 2024 (ChiCTR2400050927). The study will be conducted in accordance with the Declaration of Helsinki and informed consent will be obtained from all patients.

## Discussion

In this randomized controlled trial, we will enroll a total of 126 unilateral ankle fracture (uni/bi-/triple ankle) incision and reduction internal fixation (ORIF) surgically fragile elderly patients to investigate the effect of ultrasound-guided high ankle block and its effect on postoperative cognitive function in such patients. In addition, we will compare the two anesthetic regimens for postoperative MoCA scores, VAS pain scores, patient satisfaction scores, first time out of bed, time to discharge and perioperative safety outcomes.

dNCR is a kind of PND, which is a kind of brain dysfunction characterized by the decline of cognitive functions such as learning, memory, mood, emotion and judgment after surgery, which can prolong hospitalization time, increase medical costs, and even develop into long-term neurocognitive disorders, which seriously affects the prognosis of the elderly patients, and brings a heavy burden to the family and the society. Foot and ankle fracture occurs in elderly patients, and elderly patients with reduced function of organs, often combined with various medical diseases, poor tolerance to anesthesia and surgery, often with various postoperative complications. It is well known that there are many factors inducing PND, and anesthesia is one of the important factors[22]. Intravertebral anesthesia is often used for lower limb fracture surgery, but elderly patients are often associated with lumbar disc herniation, ligament calcification and other problems, and are prone to urinary retention, postoperative headache and other adverse reactions, which makes it impossible to carry out intravertebral anesthesia. Simple general anesthesia due to the gradual deterioration of the function of various organs in elderly patients, reduced immunity, vascular elasticity and poor tolerance to anesthesia, coupled with the traumatic stimulation of the operation, often resulting in a pronounced stress reaction, and even cause perioperative cognitive dysfunction, which is not conducive to their prognosis [6], especially for the role of fragile elderly patients more so. According to a preliminary study presented at the annual meeting of the American Society of Regional Anesthesia and Pain Medicine in 2023 [11], blockade of the tibial, superficial peroneal, and deep peroneal, peroneal, and saphenous nerves in the middle of the calf between the popliteal fossa and the ankle provides prolonged analgesia for foot and ankle surgery while preserving ankle joint motor function. Therefore, ultrasound-guided high ankle block combined with general anesthesia was chosen in this study to improve the intraoperative safety and comfort of the patients and to improve their postoperative cognitive function. At the same time, perfect analgesia is beneficial to patients’ early postoperative functional exercise and promotes the recovery of joint function.

This experimental neuropsychological test battery consisted of simple tests covering seven different cognitive domains, and the number of test items and the cognitive domains covered conformed to the criteria recommended by the International Study Group on Postoperative Cognitive Dysfunction. The scales consisted of the Connectedness Test (Form A), the Nail Board Test (Habitual Hand), the Corsi Block Test, the Digit Symbol Replacement Subtest of the Wechsler Intelligence Scale, the Digit Breadth (Reversal) Subtest of the Wechsler Memory Scale, the Paired Associated Language Subtest, and the Visual Regeneration Subtest. The Z-value of each cognitive test was calculated according to the Z-value composite score formula for each patient. Using the 1-day preoperative test results as the baseline and comparing the differences from the baseline of each patient’s individual test results at 1 day, 7 days, and 1 month postoperatively, the Z-values for each test at the 1-day, 7-day, and 1-month postoperative time points for that patient could be calculated. The diagnosis of dNCR was made in patients in whom participating individuals had Z scores greater than 1.96 on 2 or more of the 7 cognitive tests.

EEG signals were acquired with a 64-channel EEG AC electrode system (sampling rate: 1000 Hz) Preprocessing of the EEG signals was performed using the following EEG signals: sampling, band-pass filtering from 0.2-47 Hz, EEG cut segmentation, Independent Component Analysis (ICA) for ophthalmoscopic and muscle artifact removal, and Common Averaging Reference (CAR). ApEn values were calculated for each segment, each channel, and each frequency band using MATLAB software. First, ApEn values were calculated for each channel and each segment in the following frequency bands: delta (2-4 Hz), theta (4-8 Hz), alpha 1 (8-11 Hz), alpha 2 (11-13 Hz), beta 1 (13-20 Hz), beta 2 (20-30 Hz), and gamma (30-45 Hz). Finally, for each EEG recording, these values were averaged between segments to obtain individual ApEn values for each channel in each frequency band. In order to compare the results obtained in this study with those in the literature, the same procedure was used to calculate the ApEn values in the total frequency spectrum (0.2-47 Hz) [23, 24].

The study used a combination of neuropsychological test question sets and EEG to assess the occurrence of postoperative dNCR in the 2 groups of patients, avoiding subjective bias due to the use of scales alone and improving the accuracy of the results of the study. However, there are still some shortcomings in this study: only the occurrence of postoperative dNCR in fragile elderly patients in our hospital was investigated, so the conclusion may not be generalizable and can not be extended to all age groups; nowadays, the emphasis is on comfortable medical treatment, so this study should prolong the postoperative follow-up time to evaluate the long-term prognosis of the patients; there is no other experimental study on the effect of ultrasound-guided high ankle block on postoperative cognitive function, so whether the ultrasound-guided high ankle block is able to reduce the incidence of postoperative dNCR effectively. Whether ultrasound-guided high ankle block can effectively reduce the incidence of postoperative dNCR needs to be further explored.

In conclusion, this randomized controlled trial will determine the effect of ultrasound-guided high ankle block on postoperative cognitive function, postoperative pain, and first time out of bed in surgically fragile elderly patients undergoing unilateral ankle fracture (uni/bi-/triple-ankle) incision and reduction internal fixation (ORIF). The results of this study will provide new insights into improving postoperative cognitive function in vulnerable elderly patients undergoing foot and ankle surgery.

## Data Availability

All data produced in the present study are available upon reasonable request to the authors

## Author Contributions

Ziwei Xia: schematic design, essay writing, form preparation. Guangkuo Ma: collection of information, organizing the literature. Huanjia Xue: collection of information, organizing the literature. Liwei Wang: dissertation guidance. Kai Wang: dissertation guidance, proposing the topic, thesis revision.

## Acknowledgements

The authors would like to thank the parents participating in this trial and the medical, nursing and research team for their help in study set-up, recruitment, data collection and monitoring of study data.

## Provenance and peer review

Not commissioned; externally peer reviewed.

## Acknowledgement

This work was supported by the Department of Anesthesiology of Xuzhou Central Hospital. The authors would like to thank all the guest editors and anonymous reviewers for their constructive comments.

## Funding

This work was supported by National Natural Science Foundation of China, 81700078; Xuzhou Science and Technology Plan Project, KC21055; Xuzhou Medical Key Talent Project, XWRCHT20220051.

## Disclaimer

The funding bodies have no input into the design of the study and are not involved in the preparation or submission of the manuscript for publication.

## Declaration of conffict of interest

None.

## Author Approval

All authors have read and approved the manuscript.

## Notes

### Competing Interest Statement

The authors have declared no competing interest.

### Clinical Trial

ChiCTR2400050927

